# Agreement between an artificial intelligence video-analysis platform and human evaluators scoring emergency medical technician psychomotor skills: a fully crossed reliability study

**DOI:** 10.64898/2026.08.26.26361437

**Authors:** Joel H. Otte, Autumn Cartagena

**Affiliations:** Emergency Medical Services and Fire Science Education, Des Moines Area Community College, West Des Moines, Iowa, USA; Institutional Effectiveness Department, Des Moines Area Community College, Ankeny, Iowa, USA

**Author notes:** Corresponding author: Joel H. Otte, Des Moines Area Community College, DMACC West Campus, 5959 Grand Avenue, West Des Moines, IA 50265, USA. **Preprint version note.** This is version 2 of a preprint first posted August 31, 2026 under the title “AI Video Analysis of Psychomotor Performance in EMS Education: Agreement With Human Evaluators Across Three Skills.” Version 2 is the version submitted for peer review. Relative to version 1 it corrects the reported range of differences in evaluators’ mean awarded scores (version 1 stated 13–18 percentage points on each skill; the spreads are 15, 8, and 18 points for the three skills), corrects the description of EMT-level psychomotor verification policy, and has been condensed and retitled. This article is a preprint and has not been peer reviewed.

**Keywords:** artificial intelligence, educational measurement, clinical competence, emergency medical technicians, observer variation, reproducibility of results

## Abstract

**Objectives:** Psychomotor competency verification for emergency medical technician (EMT) candidates is set by each state and, where states approve program-verified competency, rests on programs and their local evaluators. Commercial artificial intelligence (AI) video-analysis platforms are marketed to programs, but none has been evaluated against human evaluators on EMT skills. We measured the inter-rater reliability of certified evaluators scoring identical video-recorded performances, then evaluated one platform against that baseline.

**Methods:** Five human evaluators and one AI platform independently scored every video of three skills in a fully crossed design: cervical collar application (15 performances), bag-valve-mask ventilation (14), and medical patient assessment (15), using identical dichotomous checklists with critical-failure criteria. We computed item-level Fleiss κ, Krippendorff α, and Gwet AC1; absolute-agreement intraclass correlation coefficients (ICC) for percentage scores; and κ for critical-failure and pass/fail decisions, for the human panel alone and with the platform as a sixth rater.

**Results:** Human item-level agreement was moderate on all three skills (κ 0.41–0.47). Single-rater score reliability was moderate (ICC 0.54–0.69) against 0.85–0.92 for the five-rater mean. Agreement on an objective 75% threshold was moderate (κ 0.54–0.57), but agreement on the recorded pass/fail decision was fair (κ 0.30–0.39) and on the critical-failure determination near zero for cervical collar (0.03) and medical assessment (0.12). Platform alignment tracked rubric observability, not task complexity: correlation with the five-human mean was 0.857 for cervical collar, exceeding every human evaluator, −0.173 for bag-valve-mask, and 0.664 for medical assessment. Adding the platform improved panel reliability for cervical collar and reduced it for the others.

**Conclusions:** The platform rated a discrete, visually observable skill as well as the panel it joined and degraded where credit depended on rates, volumes, or verbalization. Formative practice feedback and adjunct rating are defensible now; autonomous summative use is not. The human baseline is least reliable precisely where it decides outcomes.

## Introduction

Psychomotor instruction and competency verification are the most instructor-hour-intensive activities an emergency medical services (EMS) program undertakes. Both require a qualified evaluator to observe each student directly, often repeatedly, at low student-to-instructor ratios, and programs recruit laboratory faculty from the same strained workforce they exist to replenish (“Addressing the EMS Workforce Shortage,” 2025; PSG Learning, 2026). That burden is shifting toward programs. At the advanced life support levels, the National Registry discontinued its psychomotor examination after June 30, 2024 in favor of program-verified student minimum competency (National Registry of Emergency Medical Technicians [NREMT], 2023, n.d.-b; “NREMT Announces Date,” 2024); at the emergency medical technician (EMT) level, the psychomotor process is set by each state (NREMT, n.d.-e), and where a state approves program-verified minimum competency in place of a state examination, hands-on verification rests almost entirely with individual programs and their local evaluator pools.

Human observation is not only expensive; it is variable. Multi-facet Rasch modeling of more than 140,000 marks in the MRCP(UK) clinical examination attributed roughly 12% of scoring variance to examiner leniency and stringency, the “hawk–dove effect,” against 1% to differences between stations (McManus et al., 2006). Differential stringency moves candidates across the pass/fail boundary (Homer, 2022), and examiner effects replicate across institutions (Schleicher et al., 2017). EMS education is not exempt: the National Registry’s own competency materials acknowledge that scoring of an identical performance varies with an evaluator’s bias, attentiveness, and command of the standard, and the checklist-with-critical-criteria format was designed in part to constrain that inconsistency (Georgia Department of Public Health, n.d.; NREMT, n.d.-c). Yet a checklist does not remove the judgment inside each checkbox, and live, unrecorded scoring leaves no record to consult when a student disputes an assessment.

Video recording creates a durable record that multiple reviewers can assess, but it relocates the bottleneck rather than removing it, because an expert must still watch the video (Pedrett et al., 2023). Artificial intelligence (AI) video analysis is the proposed remedy, most maturely in surgery (Hashemi et al., 2025; Pedrett et al., 2023; Zeng et al., 2026), increasingly in clinical skills examinations (Kang et al., 2026; León-Ariza et al., 2026), and, adjacent to EMS, in computer-vision assessment of cardiopulmonary resuscitation from ordinary camera video (Constable et al., 2025; de Raad et al., 2026; Wang et al., 2026). For a program the appeal is capacity: an automated evaluator is consistent, tireless, and available whenever the student is, permitting unlimited scored practice of the kind that improved subsequent faculty-scored performance in a randomized trial (Lavigne et al., 2025).

The evidence counsels caution in equal measure. Reviews describe a feasibility-dominated literature in which automated systems award higher and more uniform scores than humans, with agreement strongest for discrete, visually observable actions and weakest for elements requiring communication or contextual judgment (León-Ariza et al., 2026), although a large language model grading written clinical reports was, by contrast, more stringent than human graders (Luordo et al., 2025). Commercial platforms are meanwhile marketed directly to EMS programs, yet to our knowledge none has been evaluated against human evaluators on EMS psychomotor skill examinations. Such an evaluation requires the right benchmark: not perfection, but the measured reliability of the evaluators the system would supplement. This study therefore asked a single question: how does a commercially available AI video-analysis platform compare with certified human EMS evaluators scoring the same video-recorded performances at the item, score, and decision levels, measured against the inter-rater reliability of those evaluators themselves, across three skills spanning a gradient of complexity?

## Methods

### Study design

We conducted a prospective, fully crossed reliability and agreement study comparing an AI video-analysis platform with human evaluators of prehospital psychomotor skills. Video-recorded performances by EMT students were scored independently by five human evaluators and by the platform using identical skill-specific rubrics, every rater scoring every available video for each skill. The study was approved by the Des Moines Area Community College Institutional Review Board (IRB ID 202612710237), and all participants provided written informed consent. The study is reported in accordance with the Guidelines for Reporting Reliability and Agreement Studies (GRRAS; Kottner et al., 2011); the completed checklist is provided as a supplementary file.

### Setting and participants

Participants were students in an initial EMT education program at Des Moines Area Community College, near completion of the course. Participation was voluntary and had no bearing on course standing. Nineteen students consented, performing up to three skill stations in a simulation laboratory as scheduling permitted; 12 completed all three. Sample size was determined by the consenting students available in a single cohort and the recording window within the course schedule, not by a priori calculation. After the exclusions described below, the analyzable sets comprised 15 cervical collar, 14 bag-valve-mask (BVM), and 15 medical assessment performances.

### Skill stations and scoring instruments

Three skills were selected a priori to represent an escalating gradient of complexity and evaluator judgment: cervical collar application, a rudimentary and highly observable task; BVM ventilation of an apneic adult patient, combining a physical sequence with time-and rate-dependent judgments; and a full medical patient assessment, requiring integrated clinical judgment from scene size-up through verbal reporting (Table 1).

**Table 1.** Skill stations, scoring instruments, and analyzable video sets.

| Skill | Checklist items | Critical-failure criteria | Analyzable videos |
| --- | --- | --- | --- |
| Cervical collar application | 10 | 4 (e.g., BSI/PPE omission, inappropriate head/neck movement, omission of pulse–motor–sensory checks, incorrect collar sizing) | 15 |
| BVM ventilation of an apneic patient | 19 | 11 (e.g., ventilation delayed or interrupted >30 s, failure to suction before ventilating, incorrect ventilation rate, inadequate or excessive volume, failure to provide high-concentration oxygen) | 14 |
| Medical assessment | 37 scoreable items across 8 domains, plus a scenario-variable medication and oxygen-therapy section | Scenario-specific critical criteria | 15 |

*Note.* BSI/PPE = body substance isolation/personal protective equipment; BVM = bag-valve-mask.

Instruments were developed by the study team: the medical assessment checklist from the National Registry Patient Assessment/Management – Medical skill sheet, the BVM checklist more loosely from the National Registry BVM Ventilation of an Apneic Adult Patient sheet, and the cervical collar checklist de novo (NREMT, n.d.-a, n.d.-d). Students were instructed and assessed using the same rubrics. Each item was scored 1 (performed correctly) or 0 (omitted or performed incorrectly), and each instrument specified critical criteria whose violation constituted an automatic failure regardless of checklist score. For the medical assessment, scenario-dependent medication and oxygen-therapy items were scored only when applicable and the denominator adjusted accordingly. The passing standard was at least 75% of possible points and the absence of any critical failure.

### Video recording

Each performance was video recorded with one fixed, single-angle, audio-capturing high dynamic range camera. Identical, unedited files were used for both the human and AI evaluations, so that all raters had access to the same visual and auditory information.

### Human evaluator panel

Five evaluators (E1–E5), all certified EMS clinicians (two paramedics, three EMTs) active in laboratory instruction, independently scored the videos in individually distributed workbooks, without access to one another’s scores or to the AI output. The panel was deliberately constructed to span the experience range found among skill examiners: one evaluator had more than 15 years of EMS experience, one between five and ten years, one three years, and two approximately one year. For the cervical collar and BVM skills, evaluators scored each video twice: an initial one-pass score recorded while watching straight through in real time, then a revised score with unlimited pausing and rewatching, to quantify information missed in real time but recoverable on replay. One evaluator completed only the single round for the cervical collar. Unless otherwise specified, the one-pass round served as the primary human data.

### AI video analysis

The same files were uploaded to a commercially available AI video-analysis platform (Vosaic AI Mate, version 1.1.5766; Vosaic, 2026) between March and June 2026. For each video the platform was provided the skill-specific rubric and analyzed the performance in two passes, video analysis followed by a repeat analysis incorporating the session transcript, generating a structured report with item-level determinations, a total score, identified critical failures, and a pass/fail decision. Reports were transcribed into the human scoring structure. Configuration of the platform, the creation and iterative refinement of rubrics and scoring instructions using a general-purpose large language model (Claude Opus 4.8; Anthropic, 2026), was completed before study scoring began; a preliminary calibration pass during BVM configuration was excluded from all analyses. The AI was the only rater to award partial (half-point) credit, which was retained as reported.

### Data handling

Item cells left blank by an evaluator were coded as 0, a convention verified against each evaluator’s own computed percentage column. Two re-recorded cervical collar videos scored by only one evaluator and one BVM placeholder row for a video never distributed were excluded. Where an AI report contained an internal discrepancy between line-item scores and its summary total, the line-item scores were used. For the medical assessment, the AI awarded points on scenario-variable items that every human treated as not applicable; comparative analyses therefore used the 37 items common to all raters, while each rater’s recorded critical-failure and pass/fail determinations were retained as reported.

### Ethical considerations

Written informed consent was obtained from all students and evaluators before any recording or scoring. Students were informed that participation was voluntary, had no bearing on course standing or certification eligibility, and that study scores would not enter the course record; evaluators were assured that participation would not affect any employment-related decision. Because students are potentially vulnerable within the instructor–learner relationship, consent was obtained by a study team member with no grading authority over participants. Scoring sheets referenced each performance by sequential code, and the analytic dataset contains no student identifiers.

### Statistical analysis

The analysis proceeded in two stages for each skill: characterization of inter-rater reliability among the five human evaluators, establishing the panel as the reference standard, then evaluation of the AI both as an additional rater within the panel and against the human consensus. At the item level, across all rubric items and videos, we computed raw percent agreement as the mean proportion of exact matches across rater pairs, Fleiss’ κ for multiple raters (Fleiss, 1971), Krippendorff’s α (Krippendorff, 2019), and Gwet’s AC1 (Gwet, 2008); Gwet’s AC1 was included because it is robust to the prevalence paradox that deflates κ-family statistics when items are credited on most performances. At the score level, agreement on the percentage score was quantified with two-way random-effects, absolute-agreement intraclass correlation coefficients (ICC) for a single rater, ICC(2,1), and for the k-rater mean, ICC(2,k) (Shrout & Fleiss, 1979). All coefficients were then recomputed with the AI as a sixth rater to quantify whether it improves, preserves, or degrades panel reliability.

Score agreement was assessed with Pearson and Spearman correlations, and consensus alignment by correlating each rater’s scores with the mean of all remaining raters; for the AI, with the five-human mean. Decision-level agreement on the recorded pass/fail decision and the critical-failure determination was quantified with Fleiss’ κ. Because the recorded decision incorporates each rater’s subjective application of the critical-failure override, we additionally computed an objective pass indicator (score ≥75%) to separate disagreement in item scoring from disagreement in the critical-failure judgment. Decisions were also compared against the human-majority verdict, the decision of at least three of five human evaluators. For each item, the AI’s award rate was compared with the mean human award rate, and for evaluators completing both rounds every video-by-item decision was classified as unchanged, credited only on replay, or revoked on replay.

Ninety-five percent confidence intervals came from nonparametric bootstrap with 2,000 resamples of participants (Efron & Tibshirani, 1993). Coefficients were interpreted against the Landis and Koch (1977) scale, Krippendorff’s (2019) threshold of α ≥ 0.667, and published ICC guidelines (Koo & Li, 2016). Because the same students appeared across skills, analyses were conducted separately by skill with no cross-skill pooling. Analyses used Python 3 (Python Software Foundation, n.d.).

### Use of artificial intelligence

The authors wrote the manuscript. Claude (Anthropic; Claude Opus 4.8 and later 2026 releases) was used to assist with language and stylistic editing, structuring and formatting of the author-written text, organization and cleaning of the scoring data, generation of the statistical analysis code, production of the tables from the analysis output, and literature searching and reference formatting. It was not used to produce the results, discussion, or conclusions. The authors reviewed and verified all content and take full responsibility for the accuracy and integrity of this publication. The use of a large language model to develop the scoring rubrics supplied to the platform under study is a study method, described above.

## Results

Values in parentheses after coefficients are 95% bootstrap confidence intervals. Fifteen cervical collar, 14 BVM, and 15 medical assessment performances were analyzed; all five evaluators and the AI scored every video, and the initial one-pass round served as the human data throughout.

### Human inter-rater reliability: the baseline

Table 2 reports the humans-only reliability coefficients; three patterns emerged.

**Table 2.**
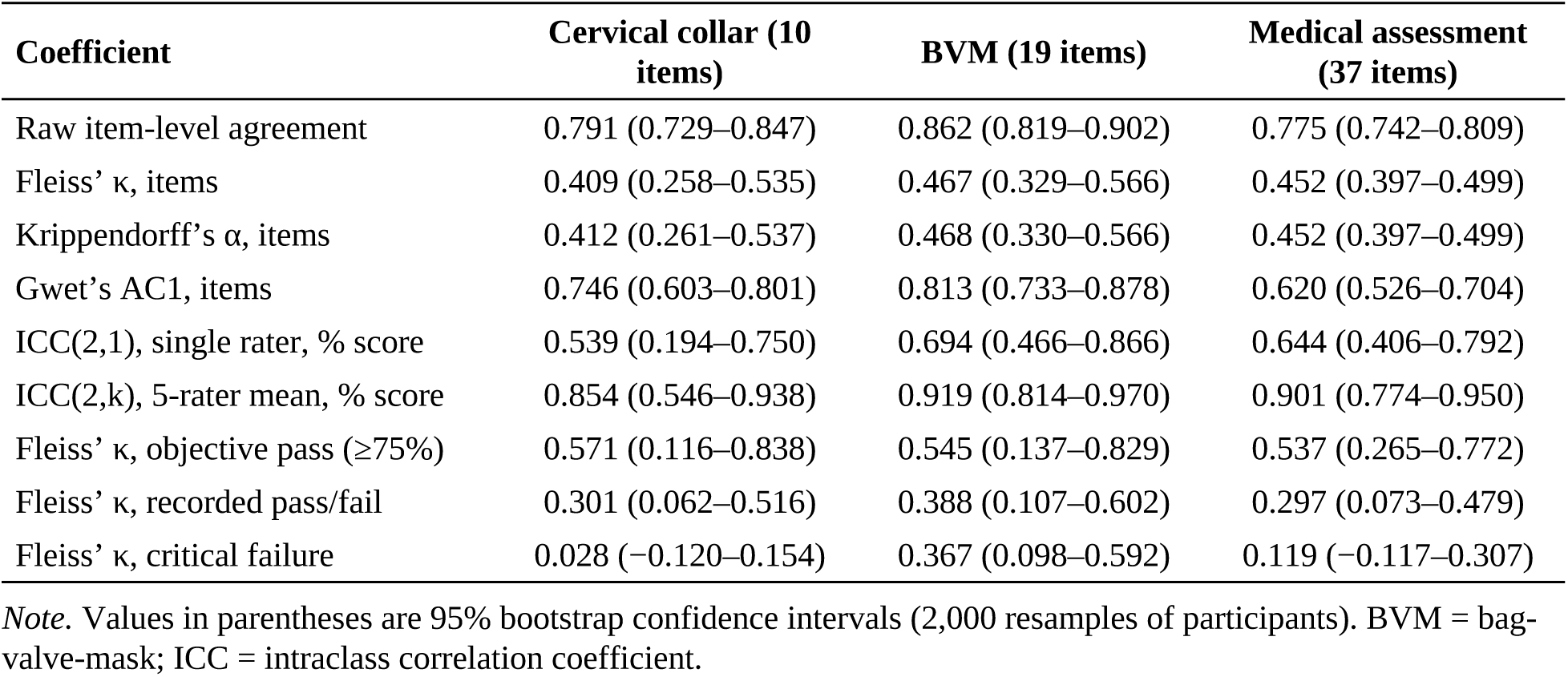
Inter-rater reliability of the five human evaluators, by skill (initial one-pass round)

First, item-level agreement among trained evaluators watching identical videos was only moderate: raw agreement ranged from 77.5% to 86.2%, and chance-corrected coefficients converged on κ ≈ α ≈ 0.41–0.47 for every skill. Agreement was highest for discrete, directly observable actions and lowest for items requiring sustained observation or judgment (Appendix A, Table A1).

Second, reliability depended strongly on whether one examiner or a panel is considered. A single evaluator’s percentage score was moderately reliable, ICC(2,1) 0.54–0.69, whereas the five-evaluator mean was good to excellent, ICC(2,k) 0.85–0.92: one examiner working alone, the usual condition in EMS testing, carried substantially more error than the panel average.

Third, and most consequentially, the pass/fail decision was markedly less reliable than the underlying scores, and the critical-failure override was the dominant source of that disagreement. Agreement on the objective pass indicator was moderate (κ 0.54–0.57), but agreement on the recorded decision fell to the fair range (κ 0.30–0.39). Agreement on the critical-failure determination itself was essentially absent for the cervical collar (κ = 0.03) and the medical assessment (κ = 0.12); only BVM, whose critical criteria are comparatively concrete, produced non-trivial agreement (κ = 0.37). The practical spread was wide (Table 3): on the cervical collar, evaluators invoked a critical failure between 0 and 13 times across 15 videos and passed between 2 and 11 students, and mean awarded scores differed between the most lenient and most severe evaluator by 15 percentage points for the cervical collar, 8 for BVM, and 18 for the medical assessment.

**Table 3.** Rater leniency and decision profiles (humans-only baseline; AI shown for reference)

| **Evaluator** | **Cervical collar: mean %**<br>**(CF; pass of 15)** | **BVM: mean % (CF; pass of**<br>**14)** | **Medical assessment: mean**<br>**% (CF; pass of 15)** |
| --- | --- | --- | --- |
| E1 | 78.7 (8; 6) | 82.7 (7; 6) | 67.4 (2; 6) |
| E2 | 70.7 (9; 5) | 80.1 (10; 4) | 78.7 (5; 8) |
| E3 | 86.0 (0; 11) | 87.2 (9; 5) | 79.8 (5; 9) |
| E4 | 73.3 (13; 2) | 85.7 (13; 1) | 62.2 (14; 1) |
| E5 | 77.0 (0; 10) | 87.6 (4; 10) | 68.3 (10; 5) |
| AI | 83.0 (5; 10) | 91.7 (8; 6) | 90.3 (10; 5) |

*Note.* CF = critical failures flagged; pass = performances marked PASS (recorded decision). Medical assessment means are computed on the 37 items common to all raters.

Unlimited review added little beyond a single real-time viewing. Among the four evaluators who completed both rounds, a full rewatch changed 19 of 600 video-by-item decisions for the cervical collar (3.2%) and 14 of 1,064 for BVM (1.3%). Approximately 97– 99% of real-time decisions survived unchanged, indicating that disagreement arose principally from interpretation rather than missed observation.

### AI performance versus the human panel

The AI was evaluated against this baseline in three ways: its effect on panel reliability as a sixth rater (Table 4), its alignment with the human consensus score (Table 5), and its item-and decision-level behavior. Item-level α used the interval metric to accommodate the AI’s half-point credit; item-level divergences between the AI and the human panel are tabulated in Appendix A (Table A2).

**Table 4.** Effect of adding the AI as a sixth rater on panel reliability.

| **Coefficient** | **Cervical collar:**<br>**humans → all six** | **BVM: humans → all**<br>**six** | **Medical assessment:**<br>**humans → all six** |
| --- | --- | --- | --- |
| ICC(2,1) | 0.539 → 0.578 | 0.694 → 0.462 | 0.644 → 0.494 |
| ICC(2,k) | 0.854 → 0.892 | 0.919 → 0.838 | 0.901 → 0.854 |
| Krippendorff's $\alpha$, items | 0.412 → 0.390 | 0.468 → 0.385 | 0.452 → 0.390 |
| Fleiss' $\kappa$, recorded pass/fail | 0.301 → 0.342 | 0.388 → 0.313 | 0.297 → 0.338 |
| Fleiss' $\kappa$, critical failure | 0.028 → 0.112 | 0.367 → 0.291 | 0.119 → 0.164 |

*Note.* BVM = bag-valve-mask; ICC = intraclass correlation coefficient.

**Table 5.**
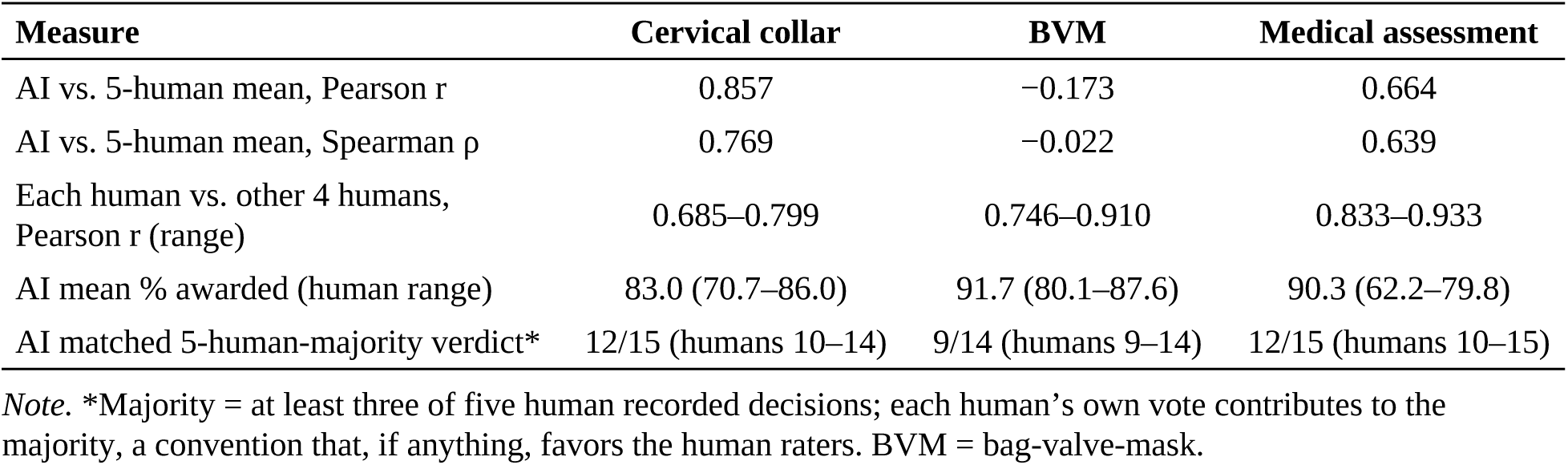
AI alignment with the human panel, by skill.

| **Measure** | **Cervical collar** | **BVM** | **Medical assessment** |
| --- | --- | --- | --- |
| AI vs. 5-human mean, Pearson $r$ | 0.857 | -0.173 | 0.664 |
| AI vs. 5-human mean, Spearman $\rho$ | 0.769 | -0.022 | 0.639 |
| Each human vs. other 4 humans,<br>Pearson $r$ (range) | 0.685–0.799 | 0.746–0.910 | 0.833–0.933 |
| AI mean % awarded (human range) | 83.0 (70.7–86.0) | 91.7 (80.1–87.6) | 90.3 (62.2–79.8) |
| AI matched 5-human-majority verdict* | 12/15 (humans 10–14) | 9/14 (humans 9–14) | 12/15 (humans 10–15) |

*Note.* \*Majority = at least three of five human recorded decisions; each human's own vote contributes to the majority, a convention that, if anything, favors the human raters. BVM = bag-valve-mask.

#### Cervical collar application

On the most basic skill the AI was the best-aligned rater on the panel, correlating with the five-human mean at r = 0.857, higher than any individual evaluator achieved against the mean of their four peers (r = 0.685–0.799). Adding it improved score-level reliability and decision-level agreement and left item-level α essentially unchanged (Table 4). Its leniency and decisions fell within the human ranges (Table 3), and it matched the human-majority verdict on 12 of 15 videos (humans 10–14). Of three discordant decisions, one was a substantive over-credit (AI 90% and pass against a human mean of 64% and a majority fail); the other two were high-scoring videos that a human majority failed through individually applied critical-failure overrides.

#### BVM ventilation

At the decision level the AI looked unremarkable: aggressive critical-failure enforcement (8 of 14 videos) pulled its pass count (6) into the human range, and it matched the human majority on 9 of 14 videos (humans 9–14). That surface normality concealed the weakest performance of the study. The AI was the most lenient and most compressed rater (mean 91.7%, standard deviation 0.09, minimum 73.7%), and, uniquely among the three skills, its scores bore no relationship to the human ranking of the same performances (r = −0.173, against human-versus-peer values of 0.746–0.910). The collapse was driven by outright inversions: on two videos the AI awarded a perfect 19/19 against failing human scores (one unanimous; human scores 11–14/19, mean 62%), while its own two lowest scores (14 and 14.5 of 19) fell on videos the humans scored 81–93%. Adding the AI lowered every reliability coefficient (Table 4).

#### Medical assessment

Against the most complex instrument the AI showed the most pronounced leniency and compression of the study. Its mean on the 37 common items was 90.3%, above the entire human range (62.2–79.8%), and its correlation with the five-human mean was positive but modest (r = 0.664), below every evaluator’s alignment with peers (0.833–0.933). The AI also scored the scenario-variable medication and oxygen-therapy sections on every performance (denominator 47), which all five evaluators treated as not applicable (denominator 37). Adding the AI reduced score-and item-level reliability while slightly raising decision-level agreement (Table 4), because its frequent critical-failure calls (10 of 15) overlapped with those of the stricter evaluators. Its over-crediting concentrated on the least observable, verbalization-dependent items (Appendix A, Table A2). Yet it matched the human majority on 12 of 15 videos (humans 10–15), and two of its three discordances were videos it failed on its own critical-failure doctrine despite awarding near-perfect scores: one at 97%, for aspirin placement and oxygen device application not visually confirmed, and one at 100%, for nitroglycerin administered without a visually confirmed blood pressure measurement.

#### Cross-skill pattern

Two signatures were consistent across skills: the AI over-credited foundational, setup, and verbalization items on reasonable presumption where humans withheld credit, and it enforced critical-failure criteria by strict visual confirmation, at times failing performances to which it had awarded near-perfect scores.

## Discussion

### Principal findings

Two findings must be read together. First, the human baseline: five trained EMS evaluators scoring identical video-recorded performances agreed only moderately at the item level (κ 0.41–0.47), a single evaluator’s percentage score was only moderately reliable (ICC 0.54–0.69), and the pass/fail decision was the least reliable output of all (κ 0.30–0.39), with the critical-failure override, nearly unreliable in itself on two of three skills, the dominant source of that disagreement. Second, against that baseline, the AI’s alignment tracked not the complexity of the task but the observability of the rubric.

### The human baseline in context

This is not an indictment of these particular evaluators; it is consistent with two decades of assessment research (Homer, 2022; McManus et al., 2006; Schleicher et al., 2017) and with what the National Registry’s own materials acknowledge (NREMT, n.d.-c). The gap between objective-threshold agreement (κ 0.54–0.57) and recorded-decision agreement (κ 0.30–0.39) localizes the problem: evaluators largely agree on what score a performance earned and disagree on whether a critical failure occurred, so the least reliable element of the examination is the one holding summative veto power. And because unlimited review changed only 1.3–3.2% of item decisions, that disagreement is interpretive rather than observational: evaluators are not missing events but construing them differently.

### Agreement with prior work

Our findings replicate, in an EMS context, the two most consistent observations in the emerging literature: automated systems award higher and more uniform scores than human raters, with agreement strongest for discrete, visually observable actions and weakest for elements requiring communication or contextual judgment (León-Ariza et al., 2026), a signature we observed on all three skills; and pose-estimation systems assess rescuer position accurately while erring on compression depth and rate (de Raad et al., 2026), a pattern our BVM findings extend to ventilation. To our knowledge this is the first published evaluation of a commercial AI video-analysis platform against human evaluators on EMS psychomotor skill examinations, and the first in any domain to benchmark one simultaneously against the item-, score-, and decision-level reliability of the human panel itself.

### Interpreting the three profiles

An observability gradient explains results that a complexity gradient cannot: performance is good wherever the thing being judged is visible in the available evidence, regardless of difficulty. The cervical collar rubric consists almost entirely of discrete, binary, visually verifiable actions, and there the AI functioned as a legitimate sixth rater. The BVM rubric embeds continuous quantities, ventilations per minute, volume adequacy, and suction duration, that are genuinely difficult to extract from ordinary video, and the AI’s response was systematic over-crediting of what it could see and misranking of what it could not. The medical assessment revealed a third failure mode, less about perception than about evaluation philosophy: the AI credited verbalization-dependent items far more liberally than the evaluators, which, given its second analytic pass over the transcript, reflects a low threshold for crediting verbal mention rather than an inability to hear it, while simultaneously enforcing critical-failure criteria by a strict visual-confirmation doctrine that failed near-perfect performances over treatment actions humans credited from context. These are not random errors but the consistent application of a different standard. The AI’s behavior thereby exposes that the human standard itself is unsettled. Whether credit requires visual confirmation or may be granted from verbalization and context is precisely the question on which the evaluators disagreed with one another, and an automated evaluator forces a program to answer it explicitly rather than leaving it to each examiner’s discretion.

### Implementation context and the improvement ceiling

That the cervical collar results reached panel-grade alignment suggests these findings describe a floor rather than a ceiling. The configuration evaluated here was an early, non-specialist implementation: a general-purpose product, not designed for EMS psychomotor examination, adapted by an EMS program director with no background in software development (Anthropic, 2026). The platform struggled to distinguish visually between airway adjuncts unique to prehospital practice, most notably an i-gel supraglottic airway from an oropharyngeal airway, a limitation plausibly connected to the BVM results. Because the platform already performs a transcript pass, the medical assessment results indicate that the need is not speech access but calibration: of the threshold at which a verbal mention earns credit, and of how transcript evidence is reconciled with visual evidence when the two diverge. The platform had transcript access throughout and still misranked BVM performances, so its failure is not the audio-deprivation failure described for visual-only grading (Kang et al., 2026). Both gaps are tuning problems on an architecture that already works.

A related observation concerns how this study was possible at all. Programs like this one sit outside the research infrastructure that funded studies assume. The roles such a study would ordinarily purchase, statistical programming, data curation, literature retrieval, and technical editing, were performed by the authors with the generative AI assistance declared in the Methods; the conception, design, execution, and interpretation were the authors’ throughout. The same scarcity of expert time that rations psychomotor instruction rations the evidence base about it, and the programs now carrying sole responsibility for competency verification are the least equipped to study what that verification is worth.

### Practical implications

The results support a differentiated rather than wholesale answer. The defensible near-term roles are those where the technology’s weaknesses are least consequential and its strengths, tirelessness, consistency, availability, and an itemized record tied to video evidence, are most valuable: formative practice feedback in open-laboratory settings, where a student can complete unlimited scored attempts without consuming evaluator hours (Lavigne et al., 2025), and archival scoring that converts every graded performance into a reviewable record. A role as an adjunct rater, with humans retaining decision authority, is supported by the cervical collar results for highly observable skills. What the results do not support is autonomous summative use: on no skill should the AI’s pass/fail verdict, and especially its critical-failure determination, currently substitute for human judgment, and for skills whose quality turns on rates, volumes, and durations even adjunct use is premature.

### Future directions

Programs are seeking relief from a constraint, and an agreement study establishes whether the technology is trustworthy enough to use, not what changes if it is. The outcomes that matter are capacities rather than coefficients: evaluator-hours consumed per cohort per skill, the enrollment at which verification becomes rate-limiting, and the practice repetitions that never happen because no qualified observer is available. A trial of unlimited AI-scored practice, re-evaluation of the same corpus after expert configuration, test–retest scoring of a fixed configuration, and evaluation of hybrid workflows would establish how much of the observed misalignment is implementation rather than architecture. In parallel, the profession faces a standard-setting task that predates the technology: defining explicitly whether, and for which items, credit requires visual confirmation, because an automated evaluator cannot inherit a standard the human evaluators have not agreed upon.

## Limitations

This study was conducted at a single program with a small convenience sample of near-completion students, and the confidence intervals around several coefficients are wide; findings should be read as estimates from a pilot-scale evaluation. There is no gold standard against which either humans or AI could be scored as accurate: all comparisons are of agreement, and the human consensus was itself demonstrably unreliable at the decision level, which constrains any claim about which rater was right. Decision-level concordance deserves particular care: on BVM the AI’s verdicts fell within the human range while its item scores were uncorrelated with the human ranking, so concordance alone is an inadequate and potentially misleading validity metric.

The AI results are specific to one commercial platform, one model version, and one non-specialist configuration, scored once per skill without assessment of run-to-run reproducibility; because scoring spanned four months, platform updates cannot be excluded. The AI’s half-point credit deviated from the binary instrument; the medical assessment’s scenario-variable section required restricting comparisons to the 37 common items; evaluators could not be blinded to student identity and could see their initial scores during the review round, so the replay change rate is a lower bound. Recording from a single fixed camera may have constrained what any video-based rater could observe: in the closest comparable study, a multimodal system achieved κ = 0.83 with three synchronized cameras but κ ≈ 0.72 with one (Kang et al., 2026).

Two protocol deviations are noted: all five evaluators scored every video, exceeding the approved minimum of one, and video submissions could not be de-identified at the point of scoring as the protocol described, although the analytic dataset contains no student identifiers. A planned expert review of the plausibility of each AI item determination was not conducted and is identified as future work.

## Conclusions

The binding constraint on psychomotor education in EMS is not knowledge of the standard; it is the supply of qualified human attention, and the movement of competency verification onto programs has increased demand for it at the moment it has become hardest to secure. This study asked how a commercial AI video-analysis platform compares with the evaluators it would supplement, measured against those evaluators’ own reliability. The answer is skill-specific and turns on observability. The platform scored every performance consistently and on a discrete, visually observable skill was the best-aligned rater on the panel. It failed where credit depended on continuous quantities that ordinary video does not carry, and it was markedly lenient where credit depended on verbalization. Those failures are specific and explicable, and because this was an early, general-purpose implementation, they likely represent a floor rather than a ceiling.

The uses within reach are those in which availability matters more than precision; summative use is a different matter, and these results do not support it. The human process the platform was measured against is least reliable exactly where it decides outcomes.

## Data Availability

All data produced in the present study are available upon reasonable request to the authors.

## Appendix A. Item-and domain-level agreement and item-level divergence

**Table A1.**
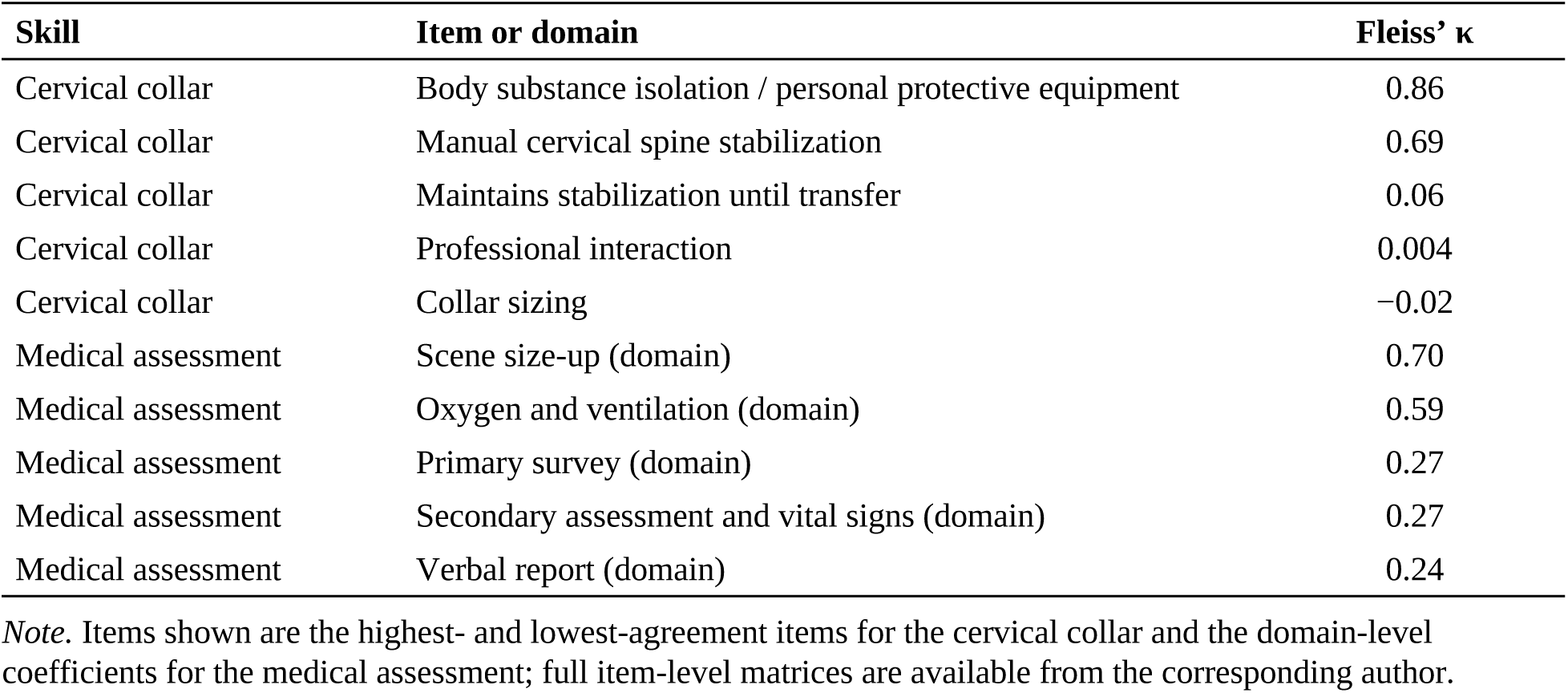
Human inter-rater agreement (Fleiss’ κ) for selected items and domains, initial one-pass round.

| Skill | Item or domain | Fleiss' $\kappa$ |
| --- | --- | --- |
| Cervical collar | Body substance isolation / personal protective equipment | 0.86 |
| Cervical collar | Manual cervical spine stabilization | 0.69 |
| Cervical collar | Maintains stabilization until transfer | 0.06 |
| Cervical collar | Professional interaction | 0.004 |
| Cervical collar | Collar sizing | -0.02 |
| Medical assessment | Scene size-up (domain) | 0.70 |
| Medical assessment | Oxygen and ventilation (domain) | 0.59 |
| Medical assessment | Primary survey (domain) | 0.27 |
| Medical assessment | Secondary assessment and vital signs (domain) | 0.27 |
| Medical assessment | Verbal report (domain) | 0.24 |

*Note.* Items shown are the highest- and lowest-agreement items for the cervical collar and the domain-level coefficients for the medical assessment; full item-level matrices are available from the corresponding author.

**Table A2.** Item-level divergence between the AI and the human panel (AI award rate minus mean human award rate)

| Skill | Item | AI – human mean award rate |
| --- | --- | --- |
| Cervical collar | Neutral head alignment | +0.23 |
| Cervical collar | Manual stabilization | +0.15 |
| Cervical collar | Pre-application pulse–motor–sensory check | +0.15 |
| Cervical collar | Post-application reassessment | -0.13 |
| Cervical collar | Maintains stabilization until transfer | -0.13 |
| BVM | Attaches BVM to oxygen and inflates reservoir (15 L/min) | +0.30 |
| BVM | Opens airway manually | +0.26 |
| BVM | Sizes oral airway correctly | +0.20 |
| BVM | Inserts oropharyngeal airway | +0.20 |
| BVM | Takes/verbalizes BSI/PPE | +0.16 |
| BVM | Suctions mouth/oropharynx $\leq 10$ s | -0.11 |
| BVM | Rigid suction catheter | -0.07 |
| Medical assessment | Spinal consideration | +0.60 |
| Medical assessment | Skin assessment | +0.59 |
| Medical assessment | Focused physical examination | +0.40 |
| Medical assessment | Trend recognition | +0.40 |
| Medical assessment | Number of patients | +0.36 |
| Medical assessment | Breathing assessment | +0.33 |

| **Skill** | **Item** | **AI – human mean**<br>**award rate** |
| --- | --- | --- |
| Medical assessment | Primary survey (domain mean) | +0.32 |
| Medical assessment | Scene size-up (domain mean) | +0.24 |

*Note.* Positive values indicate the AI credited the item more often than the human panel; negative values indicate the AI withheld credit more often. Items shown are the largest divergences in each direction per skill. BSI/PPE = body substance isolation/personal protective equipment; BVM = bag-valve-mask.

