## Supplementary material for "Agreement between an artificial intelligence video-analysis platform and human evaluators scoring emergency medical technician psychomotor skills: a fully crossed reliability study": GRRAS checklist (from another version of this preprint: version 1, 2026-08-31)

### GRRAS Checklist

*Guidelines for Reporting Reliability and Agreement Studies*

#### Full-Length Manuscript

*AI Video Analysis of Psychomotor Performance in EMS Education: Agreement With Human Evaluators Across Three Skills*

Otte JH, Cartagena A.

Checklist per Kottner J, Audigé L, Brorson S, Donner A, Gajewski BJ, Hróbjartsson A, Roberts C, Shoukri M, Streiner DL. Guidelines for reporting reliability and agreement studies (GRRAS) were proposed. *J Clin Epidemiol.* 2011;64(1):96-106.

*Page numbers refer to the full-length manuscript as submitted.*

| Section | Item | Checklist item | Where addressed | Page |
|---|---|---|---|---|
| Title / Abstract | **1** | Identify in title or abstract that interrater/intrarater reliability or agreement was investigated. | Title: “Agreement With Human Evaluators Across Three Skills.” Aims paragraph: “quantify the inter-rater reliability of experienced human EMS evaluators.” | **1** |
| Introduction | **2** | Name and describe the diagnostic or measurement device of interest explicitly. | Methods, Skill Stations and Scoring Instruments (Table 1); Methods, AI Video Analysis: Vosaic AI Mate v1.1.5766. | **4** |
| | **3** | Specify the subject population of interest. | Methods, Setting and Participants: students in an initial EMT education program nearing completion of didactic and psychomotor portions. | **4** |
| | **4** | Specify the rater population of interest (if applicable). | Methods, Human Evaluator Panel: five certified EMS providers (two paramedics, three EMTs), all active in EMS education laboratory instruction. | **5** |
| | **5** | Describe what is already known about reliability and agreement and provide a rationale for the study (if applicable). | Background, paragraphs 2–5: hawk–dove effect, differential examiner stringency, cross-institutional examiner effects, NREMT acknowledgment; rationale for an EMS-specific evaluation. | **2** |
| Methods | **6** | Explain how the sample size was chosen. State the determined number of raters, subjects/objects, and replicate observations. | Methods, Setting and Participants: sample size was determined by the number of consenting students available in a single EMT cohort and by the recording window available within the course | **4** |

| Section | Item | Checklist item | Where addressed | Page |
|---|---|---|---|---|
| | | | schedule, rather than by a priori calculation. Five human raters plus one AI rater; 15 cervical collar, 14 BVM, and 15 medical assessment performances; two scoring rounds for the collar and BVM skills. | |
| | **7** | Describe the sampling method. | Methods, Setting and Participants: voluntary convenience sample of consenting students within one cohort; participation had no bearing on course standing; students completed up to three stations as scheduling permitted. | **4** |
| | **8** | Describe the measurement/rating process (e.g. time interval between repeated measurements, availability of clinical information, blinding). | Methods, Video Recording and Human Evaluator Panel: one fixed, single-angle, audio-capturing HDR camera; identical unedited files for human and AI raters; two-round design (one-pass, then unlimited replay) for collar and BVM. Protocol Deviations: evaluators not blinded to student identity; scoring recorded on de-identified score sheets. | **5** |
| | **9** | State whether measurements/ratings were conducted independently. | Methods, Human Evaluator Panel: evaluators scored independently using individually distributed workbooks, without access to one another's scores or to the AI output. | **5** |
| | **10** | Describe the statistical analysis. | Methods, Statistical Analysis: raw percent agreement, Fleiss' κ, Krippendorff's α, Gwet's AC1, ICC(2,1) and ICC(2,k), Pearson and Spearman correlation, leave-one-out consensus alignment, human-majority comparison, and 95% bootstrap intervals from 2,000 resamples. | **6** |
| Results | **11** | State the actual number of raters and subjects/objects which were included and the number of replicate observations which were conducted. | Results, Analyzed Performances: 15 collar, 14 BVM, and 15 medical assessment performances; all five human evaluators and the AI scored every video; four evaluators completed both scoring rounds. | **7** |
| | **12** | Describe the sample characteristics of raters and subjects (e.g. training, experience). | Methods, Human Evaluator Panel: two paramedics and three EMTs, all active in laboratory instruction, | **5** |

| Section | Item | Checklist item | Where addressed | Page |
|---|---|---|---|---|
| | | | with experience spanning roughly one to more than fifteen years. Subjects: EMT students nearing course completion. | |
| | **13** | Report estimates of reliability and agreement including measures of statistical uncertainty. | Table 2: all humans-only coefficients with 95% bootstrap confidence intervals. Tables 3–5: leniency and decision profiles, effect of adding the AI, and AI alignment. | **8** |
| Discussion | **14** | Discuss the practical relevance of results. | Discussion, Practical Implications and Future Directions: formative and archival use supported; summative use not supported; evaluator-hours, cost per scored attempt, and repetitions to proficiency identified as the decisive outcomes. | **14** |
| Auxiliary material | **15** | Provide detailed results if possible (e.g. online). | Data Availability: de-identified item-level scoring matrices and analysis code available from the corresponding author on reasonable request. | **20** |
